# Investigation of a Pathogenic Inversion in *UNC13D* and Comprehensive Analysis of Chromosomal Inversions Across Diverse Datasets

**DOI:** 10.1101/2024.10.28.24315942

**Authors:** Tugce Bozkurt-Yozgatli, Ming Yin Lun, Jesse D. Bengtsson, Ugur Sezerman, Ivan K. Chinn, Zeynep Coban-Akdemir, Claudia M.B. Carvalho

**Author notes:** **Co-Corresponding authors: Zeynep Coban-Akdemir,** PhD, Assistant Professor, UTHealth School of Public Health, 1200 Pressler Street, Houston, TX 77030-3900, **Claudia M.B. Carvalho**, PhD, Assistant Investigator, Pacific Northwest Research Institute, 720 Broadway, Seattle, WA 98122.

## Abstract

Inversions are known contributors to the pathogenesis of genetic diseases. Identifying inversions poses significant challenges, making it one of the most demanding structural variants (SVs) to detect and interpret. Recent advancements in sequencing technologies and the development of publicly available SV datasets have substantially enhanced our capability to explore inversions. However, a cross-comparison in those datasets remains unexplored. In this study, we reported a proband with familial hemophagocytic lymphohistiocytosis type-3 carrying c.1389+1G>A *in trans* with NC_000017.11:75576992_75829587inv disrupting *UNC13D*, an inversion present in 0.006345% of individuals in gnomAD(v4.0). Based on this result, we investigate the features of potentially pathogenic inversions in public datasets. 98.9% of inversions are rare in gnomAD, and they disrupt 5% of protein-coding genes associated with a phenotype in OMIM. We then conducted a comparative analysis of the datasets, including gnomAD, DGV, and 1KGP, and two recent studies from the Human Genome Structural Variation Consortium revealed common and dataset-specific inversion characteristics suggesting methodology detection biases. Next, we investigated the genetic features of inversions disrupting the protein-coding genes by classifying the intersections between them into three categories. We found that most of the protein-coding genes in OMIM disrupted by inversions are associated with autosomal recessive phenotypes regardless of categories supporting the hypothesis that inversions in trans with other variants are hidden causes of monogenic diseases. This effort aims to fill the gap in our understanding of the molecular characteristics of inversions with low frequency in the population and highlight the importance of identifying them in rare disease studies.

## INTRODUCTION

Inversions are defined as a type of structural variant (SV) that refers to orientation changes in DNA segments. They can be copy-number neutral (classical/simple/balanced) with two breakpoint junctions or be part of complex genomic rearrangements (CGRs) with other copy-number variations (CNVs) [1]. The main mechanism for the formation of classical inversions has been proposed to be non-allelic homologous recombination (NAHR) between inverted repeats [2–4]. Other biological mechanisms may result in inversion formation, including DNA repair-associated events (non-homologous end joining (NHEJ), and microhomology-mediated end joining (MMEJ)) and DNA replication-associated events (*e.g.,* fork stalling and template switching) [1,5].

Inversions may have an impact on disease phenotypes, often by directly disrupting a particular gene [6]. They may occur within a gene, leading to the disease manifestation by causing the skipping of exonic regions [7]. Mor-Shaked *et al*. reported a pathogenic inversion in *PRKN*, leading to the skipping of exon 5 in individuals with early-onset Parkinson’s disease (PARK2, OMIM #600116) [7]. Alternatively, one of the inversion breakpoints can disrupt a gene and result in a disease phenotype [8]. For instance, one of the breakpoints of a 253-kb inversion mapping to intron 30 of *UNC13D* contributes to the manifestation of familial hemophagocytic lymphohistiocytosis 3 (FHL3, OMIM #60898) [9,10]. In addition to Mendelian disorders, inversions are also recognized as significant contributors to common complex disease traits [11–13] and disease prognosis [14]. Additionally, they can also play a role as genetic modifiers affecting disease phenotypes [15]. Moreover, some inversions have no direct effect on disease phenotype by themselves, but they may predispose the loci to further genomic rearrangements with pathogenic consequences [2,16] including the formation of recombinant chromosomes [1]. Inversion detection is challenging due to their balanced nature and the fact that breakpoints often map to repeats. Those features make them undetectable by comparative genomic hybridization (aCGH) and exome sequencing (ES) [17]. Although short-read whole genome sequencing (WGS) enables the detection of some inversions, it also introduces the issues of false positives and the inability to sequence breakpoint junctions in the repetitive parts of the genome [18,19]. Long-read WGS technologies, including Pacific Biosciences (PacBio) and Oxford Nanopore (ONT), single-cell template strand sequencing (Strand-seq) [20], and optical genome mapping [21] have improved our ability to detect inversions since these methodologies are more suitable to detect changes in DNA orientation including within complex repeat regions [4,22].

Published population datasets using different sequencing technologies like those in Ebert *et al.* [22], and Porubsky *et al.* [4], and publicly available databases such as Genome Aggregation Database (gnomAD) [23], The Database of Genomic Variants (DGV) [24], and 1000 Genomes Project (1KGP) [25] provide valuable resources for SV analysis. The recent release of gnomAD dataset version 4 (v4.0) includes short-read genome sequencing data from 63,046 unrelated human samples across the world [23]. The DGV dataset is derived from different methodologies such as sequencing, aCGH, and Fluorescence in situ hybridization (FISH) [24]. Byrska-Bishop *et al.* released expanded short-read WGS of 1KGP consisting of 3,202 samples, including 602 trios across diverse global populations [25]. Porubsky *et al*. [4] reported inversions from 41 human samples by integrating Strand-seq [20], haplotype-resolved *de novo* sequence assemblies generated from PacBio long-reads, and Bionano genomics single-molecule optical mapping [21]. Ebert *et al.* published 64 assembled haplotypes from 32 diverse human genomes using long-read WGS and strand-seq [22].

Here, we report a proband carrying a pathogenic inversion *in trans* with a single-nucleotide variant (SNV) affecting *UNC13D*. Then, we comprehensively compare inversions disrupting genes reported in various datasets, gnomAD (v4.0) [23], DGV (release date: 2020-02-25) [24], 1KGP (release date: 2021-10-05) [25], inversions released by Ebert *et al.* [22] and Porubsky *et al.* [4] (Figure 1). Our goal is to provide insights into the features of inversions present in population datasets to genomic disorders.

**Figure 1.**
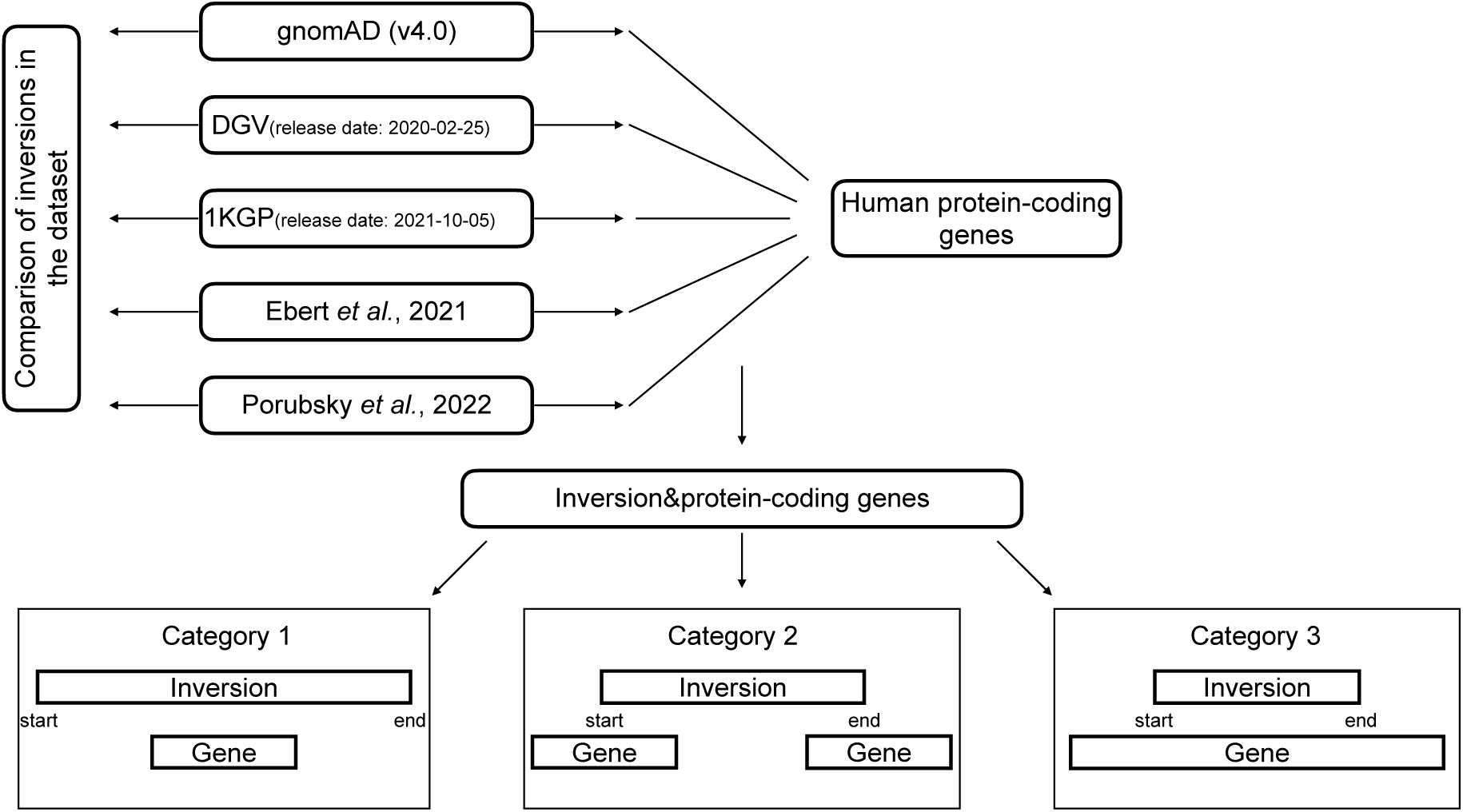
Overview of the datasets and the study design. We extracted inversions from publicly available databases, gnomAD (v4.0) [23], DGV (release date: 2020-02-25) [24], 1KGP (release date: 2021-10-05) [25] and two recent publications of Ebert *et al.* [22] and Porubsky *et al*. [4] We then intersect inversions with OMIM genes and grouped inversion-gene intersections into three categories.

## METHODS

### Case presentation

The proband (SEA110) is a Caucasian white non-Hispanic, non-Latino male. He was diagnosed with VACTERL (vertebral defects, anal atresia, cardiac defects, tracheoesophageal fistula, renal anomalies, and limb abnormalities) after birth. He did not meet early developmental milestones on time. The patient had frequent respiratory infections that required supplemental oxygen, including respiratory syncytial virus infection. He presented with pancytopenia at the range of 11-15 months of life, which was initially felt to be likely viral-mediated. He was hospitalized and discharged. He seemed well but then developed daily fevers and increased stool output. He was re-hospitalized and found to have hepatosplenomegaly by abdominal ultrasound. He then developed acute respiratory failure and required intubation with pressor support. Laboratory testing ultimately confirmed a diagnosis of hemophagocytic lymphohistiocytosis (HLH) by HLH-2004 criteria [26]: fever, splenomegaly, anemia with thrombocytopenia, hypofibrinogenemia, hypertriglyceridemia, hyperferritinemia, elevated soluble interleukin-2 receptor levels, and impaired CD107A mobilization. Further clinical information can be provided upon request from the corresponding author. Initial genetic testing consisted of proband ES and chromosomal microarray testing, both of which were performed by a commercial clinical laboratory. Results were reported as negative for both tests. Upon re-hospitalization, clinical targeted gene panel testing was ordered for inborn errors of immunity and cytopenias, which identified a pathogenic variant at *UNC13D* c.1389+1G>A.

### Patient sample collection

As a result of clinical targeted gene panel findings, SEA110 was tested by the Baylor Genetics Clinical Diagnostic Laboratory using rapid short-read WGS. Informed consent was obtained for research participation under Pacific Northwest Research Institute approved WCG IRB Protocol #H-47127_20202158.

### DNA Extraction

DNA was extracted from whole blood using the QIAGEN Puregen DNAeasy kit following the manufacturer’s direction with modification of the centrifugation steps, which were extended to 10 minutes. Ultrahigh molecular weight DNA was extracted from whole blood with the Bionano SP-G2 Blood and CellCulture DNA Isolation Kit (#80060) following the manufacturer’s direction.

### ONT-library preparation and sequencing run

DNA from SEA110 was sheared to an N50 of approximately 10 kb using a Covaris g-TUBE and an Eppendorf 5424 rotor at 5000 rpm. End repair and ligation of adapters for Oxford nanopore sequencing followed the manufacturer’s direction for kit LSK114. Sequencing used Minknow version 23.07.12, with adaptive sampling to enrich for the region of interest. The enrichment region (chr17:75526717-75896404, GRCh38) and reference as a minimap2 index file were provided [27]. Following sequencing, passed reads were re-called using guppy 6.0.1 and the super high accuracy model. Passed reads were mapped to GRCh38 using minimap2 (-Y -- secondary=no -a -x map-ont). After mapping, SNVs were called using Clair3 [28] and reads were haplotagged by Whatshap [29].

### Breakpoint junction amplification and Sanger sequencing

Inversion junctions were amplified using primers reported previously with one additional sequencing primer (Supplementary Table 1) [9]. Amplification used the Q5 Polymerase (NEB), and PCR products were gel extracted with the Monarch DNA Gel Extraction kit (NEB) following the manufacturer’s direction. Purified products were sent for Sanger sequencing by GENEWIZ. Sanger sequencing was analyzed using Geneious Prime software (Dotmatics).

### Optical Genome Mapping

Ultrahigh molecular weight DNA (UHMW) was labeled with the Bionano Direct Label and Stain-G2 (DLS2-G2) Kit (#80046) following the manufacturer’s direction. In brief, 750 ng of UHMW DNA was labeled with proprietary green fluorophore (DL-Green), and after purification, the DNA back bone was stained with a proprietary DNA stain. After staining, the sample was run on a Bionano Saphyr instrument. A *de novo* assembly was generated in Bionano access version 1.8.1, with a molecule N50 of 150.38 kb in length and 15.61 labels per 100 kb. The resulting assembly was compared to the hg38 reference genome, variants were called using Bionano solve version 1.8.1.

### Datasets utilized in this study

We analyzed the inversions mapped to the reference human genome of hg38 from three publicly accessible databases, gnomAD (v4.0) [23], DGV (release date: 2020-02-25) [24] and 1KGP (release date: 2021-10-05) [25], and two recent studies of Ebert *et al.* [22] and Porubsky *et al*. [4] (Figure 1). We extracted inversion calls in autosome (chr1-22) and sex (chrX and chrY) chromosomes from the datasets. The gnomAD (v4.0) [23] SV dataset was downloaded from https://gnomad.broadinstitute.org/downloads. The DGV [24] SV dataset was downloaded from the link: http://dgv.tcag.ca/dgv/docs/GRCh38_hg38_variants_2020-02-25.txt. DGV [24] includes inversions from several studies (Supplementary table 2) derived from different methodologies, including sequencing, oligo aCGH, and FISH. We included inversions detected by all of these studies. SV data in the 1KGP was downloaded from the following link: https://www.internationalgenome.org/data-portal/data-collection/30x-grch38. The updated callset to the original release of the inversions reported by Ebert *et al.* [22] was downloaded from the following link: http://ftp.1000genomes.ebi.ac.uk/vol1/ftp/data_collections/HGSVC2/release/v2.0/integrated_callset/. Lastly, we included the inversions reported by Porubsky *et al.* [4].

### Gene Annotations

We downloaded the gene regions with their canonical transcripts present in the hg38 version of the GENCODE (v46) database (Data update date: 2024-04-02) through the University of California Santa Cruz (UCSC) [30] to identify the inversions intersecting with the human protein-coding genes. We filtered the dataset to extract only the genes with protein-coding transcripts, excluding those with other transcript types. (Supplementary figure 1). Then, we retained the genes in human autosome chromosomes (chr1-22) and sex chromosomes (chrX and chrY). We also downloaded the dataset of the Online Mendelian Inheritance in Man (OMIM) (data freeze date: 06-18-2024) [31] (https://www.omim.org/downloads) as well as rare disease-related genes in Orphanet data (https://www.orphadata.com/genes/).

### Analysis of inversions intersecting inversions in other datasets and protein-coding genes

We used the Bedtools (v2.30.0) [32] intersect function with the fraction option 0.5 to detect the overlap between inversion locations in different datasets. Bedtools intersect function takes a genomic feature as the first input and finds overlapped regions between another genomic feature as the second input. The fraction option 0.5 allows us to find the overlap, including at least 50% of the sequence length of inversions. We also implemented the Bedtools (v2.30.0) [32] intersect function with the default parameters to detect the overlap between inversions and protein-coding genes. The intersections between inversions and human protein-coding genes were classified into three distinct categories. In category 1, inversions cover genes; in category 2, one of the inversion breakpoints maps within a gene; in category 3, inversions map entirely within genes (Figure 1).

### Enrichment analysis of the genes intersecting inversions

We performed gene set enrichment analysis with the protein-coding genes overlapping with inversions in categories 2 and 3 by applying Enrichr [33]. The list of the genes intersecting inversions in each intersection category was given as input to Enrichr [33]. Then, we reported the Human Phenotype Ontology (HPO) terms enriched by these genes.

### Computational Analysis

Computational analyses were carried out using R (v.4.2.0) [34]. The plots were generated using the package ggplot2 [35] and the UpSet R package [36].

## RESULTS

### A pathogenic *UNC13D* inversion is present in gnomAD

We identified an inversion accompanied by the canonical donor splice site SNV in *UNC13D* in SEA110 (Figure 2 and Supplementary figure 2). The 253-kb inversion has been documented in individuals with Swedish ancestry and reported to cause FHL3 when inherited as homozygous or *in trans* with pathogenic SNVs and small indels in *UNC13D* [9,10]. We observed an almost identical inversion reported in gnomAD at coordinates chr17:75576924-75829482 (INV_CHR17_66182818), which is present in 0.006345%, exclusively in heterozygous state in individuals from European Finnish and Admixed American populations (Supplementary Figure 3). The SEA110 inversion shows two breakpoint junctions with 111 (junction 1) and 23 (junction 2) nucleotides similarity generated by *Alu*-*Alu* mediated rearrangement (AAMR) (Figure 2). Parental samples are not available to test for inheritance; therefore, we do not have information about ancestry and cannot investigate whether this inversion is the same reported in gnomAD (a potential founder event) or if it is a recurrent inversion generated independently via AAMR in this proband. Optical Genome Mapping supports the breakpoint junctions of the inversion obtained by Sanger sequencing. The detected inversion has multiple molecules spanning both breakpoints and several molecules spanning the entire inversion supporting the inversion call. Bionano solve software called the inversion as heterozygous, but lack of label density in *UNC13D* results in the exclusion of *UNC13D* from the called inversion. ONT sequencing was applied to confirm heterozygosity, and manual phasing indicated the pathogenic SNV and inversions are *in trans* (Figure 2C).

**Figure 2.**
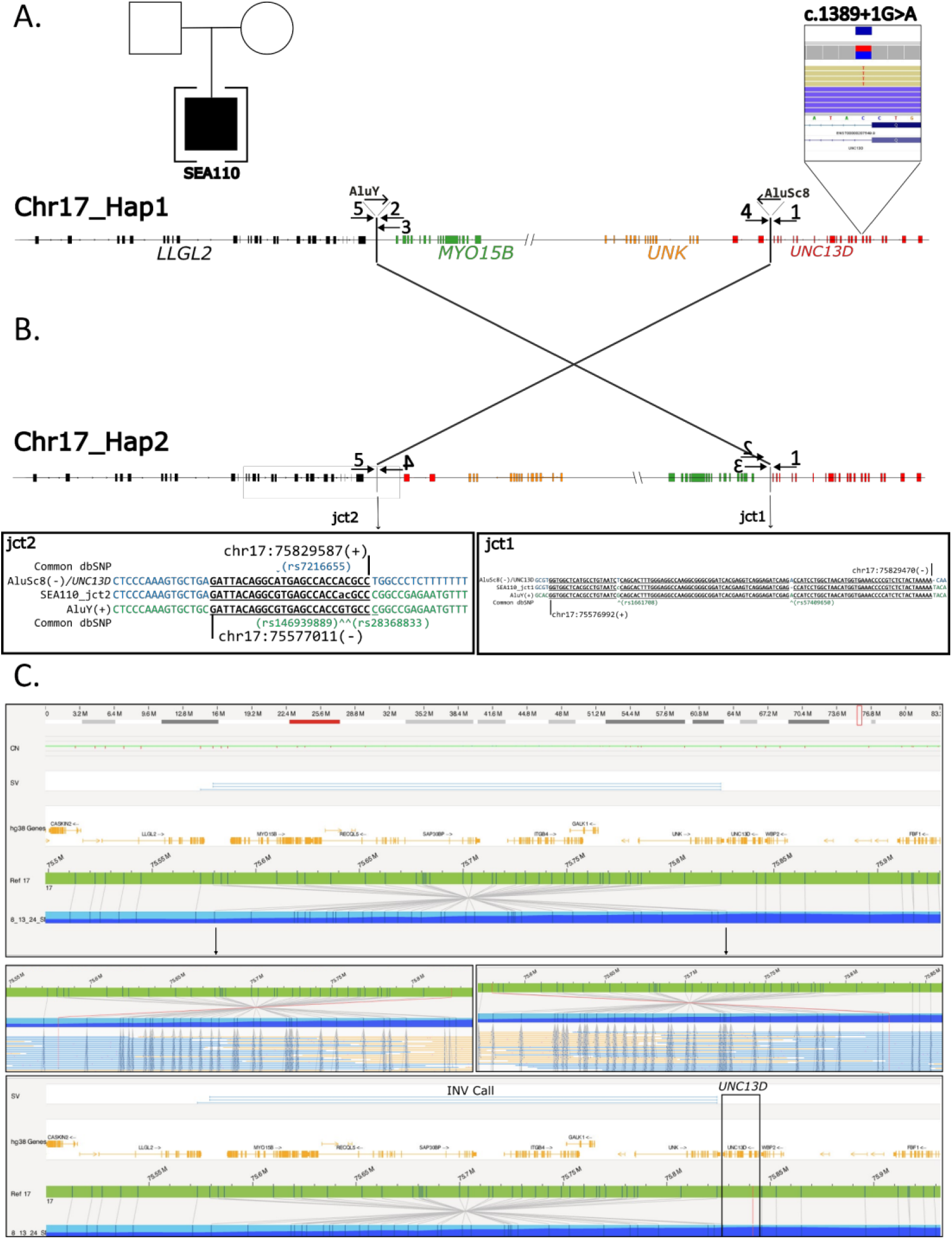
*UNC13D* variants in the patient. (A) Pedigree of patient SEA110 and IGV screenshot displaying nanopore sequencing reads that detected the pathogenic SNV in *UNC13D* (NM_199242). Chr17_Hap1 represents the haplotype carrying the SNV in *UNC13D*, the blowout of *UNC13D* point to the approximate location of the SNV. (B) Diagram of Chr17_Hap2, showing the inversion junction sequencing alignments of each breakpoint. Arrows point to the alignments for junctions 1 and 2 (jct1/2). PCR primers used to obtain the breakpoint junctions for Sanger sequencing are indicated by arrows. Arrows are not to scale. (C) Optical Genome Mapping showing the inversion in Chr17_Hap2, panels show molecules spanning each junction and the location of *UNC13D* relative to the inversion call.

### Inversions in gnomAD (v4.0) are rare and affect protein-coding genes

We hypothesized that pathogenic inversions are present as rare alleles in the general population. To investigate this concept, we categorized 2185 inversions in gnomAD into two groups: Rare (allele frequency <5%) and common (allele frequency ≥5%). Altogether, 2,161 (98.9%) inversions are rare; 24 inversions (1.1%) are common in gnomAD (Supplementary figure 4A).

We investigated the human protein-coding genes affected by rare and common inversions in gnomAD. We analyzed 19,697 protein-coding genes in GENCODE (v46); 4,921 are related to a phenotype in OMIM, 11,306 are not yet linked with a phenotype in OMIM, and 3,470 genes are not cataloged in OMIM. We overlapped inversions in gnomAD and protein-coding genes and categorized the intersections into three groups (Category 1, category 2, and category 3). Next, we focused on the inversions in categories 2 and 3 since they can be critical mechanisms for disease pathology (Supplementary table 3). 279 rare gnomAD inversions affect 5% of genes associated with a phenotype in OMIM (247 out of 4,921; Supplementary figure 4C) in contrast with 4.6% of genes not associated with a phenotype in OMIM (521 out of 11,306; Supplementary Figure 4C) based on categories 2 and 3. Furthermore, 254 out of 279 rare gnomAD inversions have not been found in the homozygous state and affect 106 autosomal recessive (AR) disease genes (Supplementary table 4).

### Features of the inversions reported in distinct datasets

To compare the characteristics of inversions in gnomAD [23] with other publicly available datasets, we conducted a comparative analysis using inversion data from DGV [24], 1KGP [25], and two recent publications of Ebert *et al.* [22] and Porubsky *et al.* [4] (Figure 1).

We extracted 2,185 inversions from gnomAD, 3,468 inversions from DGV, 920 inversions from 1KGP, 414 inversions from the data released by Ebert et *al*., and 339 inversions from the callset published by Porubsky *et al.* The summary statistics of inversion length in each dataset are provided in Table 1. gnomAD shows a more even distribution regarding size and displays the largest events (Supplementary figure 5), including a 118.67 Mb pericentric inversion (INV_CHR5_77480914). Most of DGV inversions (75%) are between 0.035 kb and 24.22 kb. 1KGP inversions tend to be smaller as the median length of 0.831 kb, whereas Ebert *et al.* and Porubsky *et al.* show the highest median length of 293.19 kb and 251.71 kb, respectively.

**Table 1.**
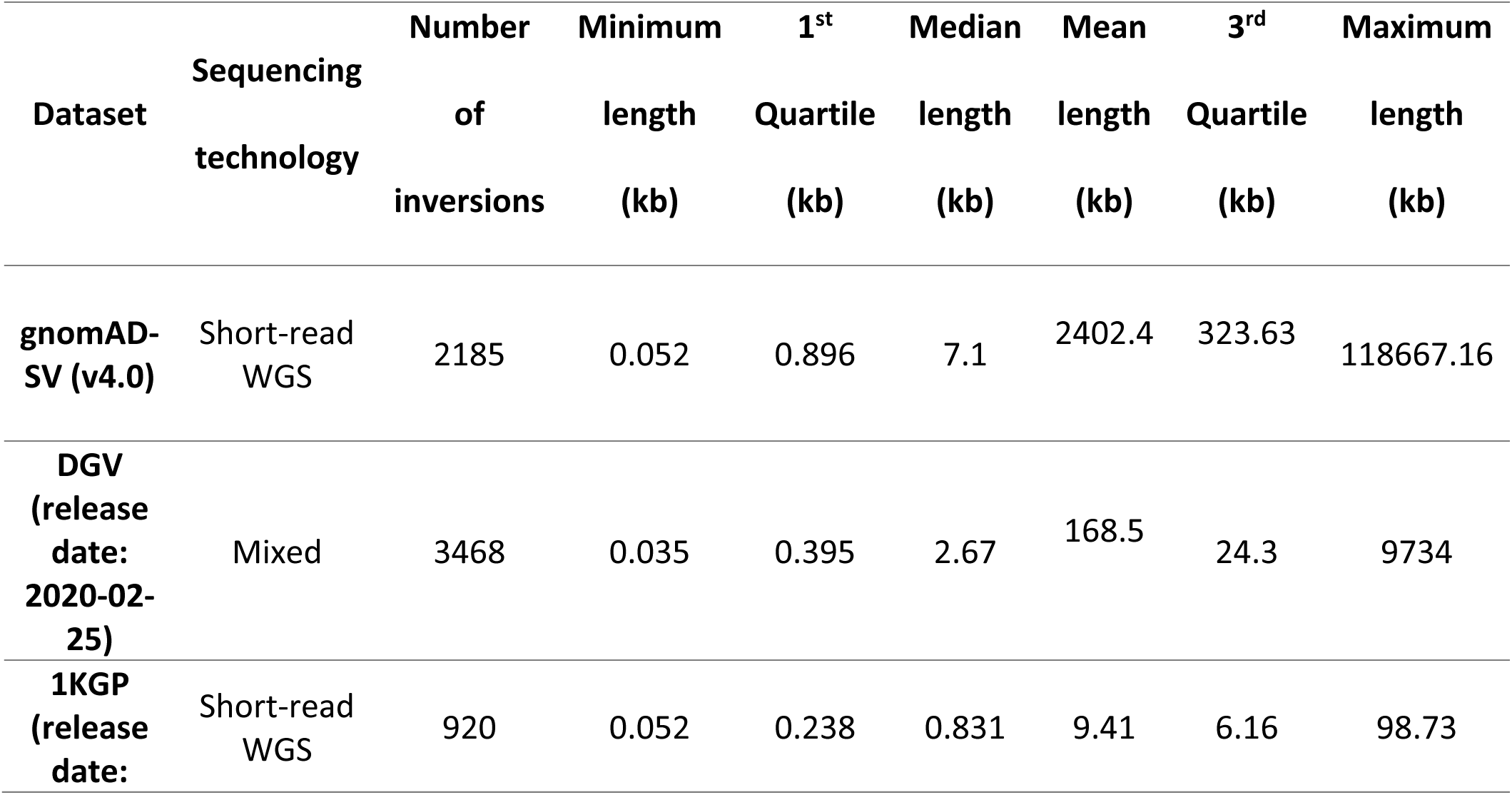
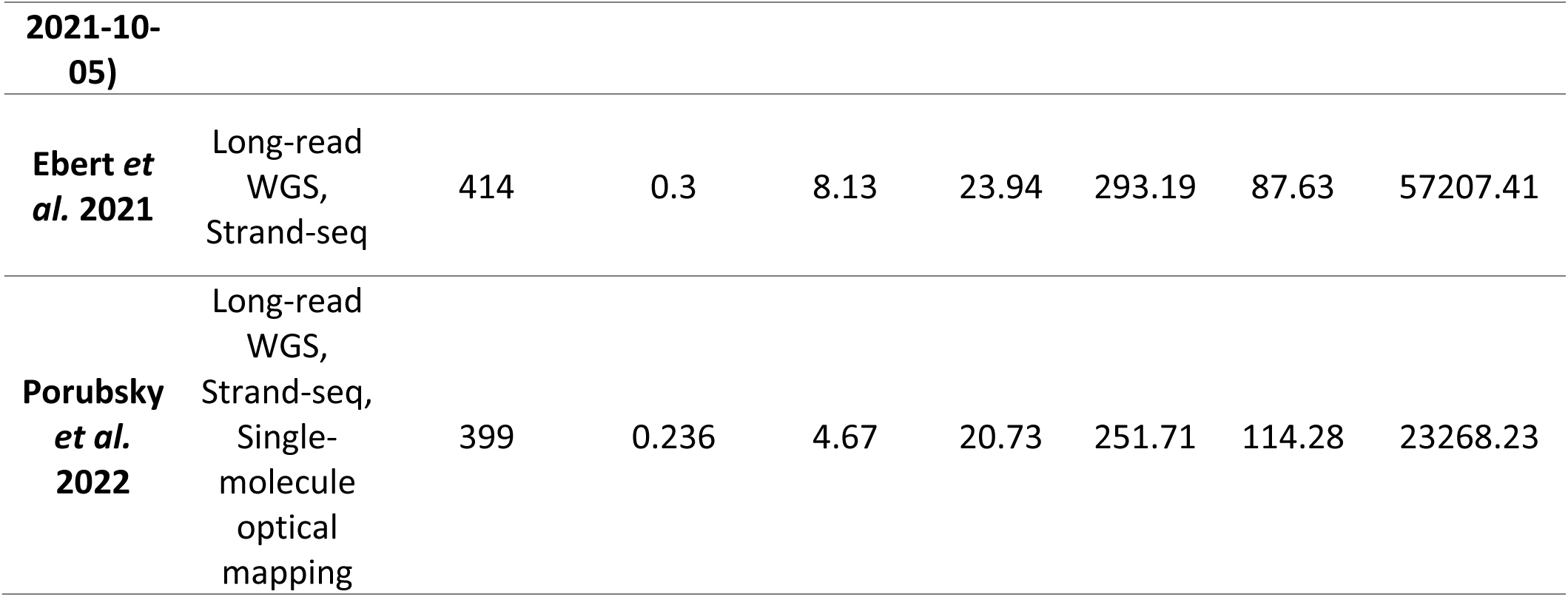
Summary statistics of the datasets analyzed in this study.

### Estimating redundancy among the inversions available from different datasets

We investigated the number of common and dataset-specific inversions across different datasets using very stringent criteria based on the start and end locations of the inversions (Supplementary figure 6). Redundancies in the datasets are expected due to the overlap of samples reported in distinct publications (*e.g.,* Ebert *et al*. and Porubsky *et al.)* or inclusion of datasets into publicly shared ones (*e.g*., gnomAD v.2 is included in DGV). We observed very little redundancy for inversions among the individual datasets (Supplementary figure 6) because the different applied sequencing technologies provided distinct resolutions concerning breakpoint junctions. We then decreased the stringency to intersect inversions in each dataset with at least 50% of their sequence (Supplementary figure 7). The inversions in gnomAD and DGV share (49.4% and 77.2%) more inversions with each other compared to other datasets. 78.3% of 1KGP inversions overlap with at least one inversion in gnomAD. Around 70% of inversions in Ebert *et al.* and Porubsky *et al.* overlap with each other.

### Inversions disrupting genes

We overlapped the inversions in the datasets with the protein-coding genes. Then, we classified the overlaps between inversions and protein-coding genes into three categories, as defined previously defined in this manuscript (Figure 1). The majority of the overlaps from all datasets, except 1KGP, map with category 1 (76.8% in DGV to 97.1% in gnomAD). 65.9% of inversion-gene intersections belong to category 3 in 1KGP (Figure 3).

**Figure 3.**
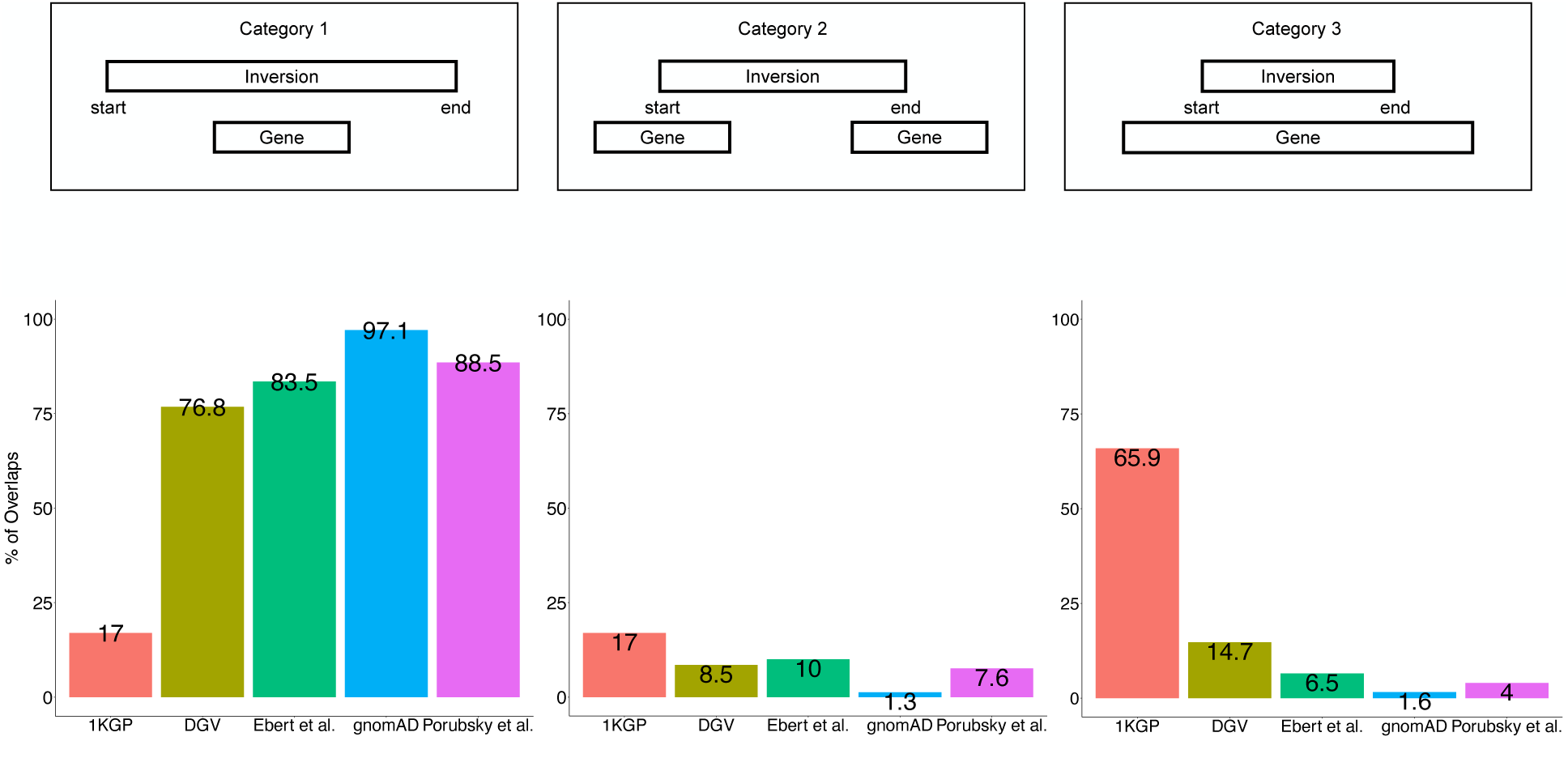
The categories of the intersections between inversions and protein-coding genes and percentages of intersections belonging to these categories. We grouped the intersections between inversions and OMIM phenotype-related genes into three categories. The first category comprises genes covered by inversions, the second category includes intersections where one of the inversion breakpoints is located within a gene region, and the third category involves inversions occurring within a gene region.

Next, we focused on the inversions in categories 2 and 3 since they can be critical mechanisms for disease pathology (Supplementary table 5 and 6). We delved deep into the protein-coding genes associated with clinical phenotypes in OMIM disrupted by inversions in this intersection between categories 2 and 3. In total, 847 inversions have one breakpoint junction mapping to 830 protein-coding genes based on category 2 and can be potentially relevant to genetic disorders (Supplementary table 5). On the other hand, in total, breakpoint junctions of 1,586 inversions are within 1030 protein-coding genes based on category 3 and can also be potentially relevant to genetic disorders (Supplementary table 6). Interestingly, both DGV and gnomAD inversions show higher frequencies of disrupting genes associated with disease compared to other datasets (1.6% and 2.1%, respectively) in Category 2 (Supplementary figure 8A). Importantly, inversions in both datasets also disrupt a higher proportion of OMIM phenotype-related genes in Category 3 (3.7% and 3.2%, respectively), while Porubsky *et al.* has a smaller proportion (0.2%, Supplementary figure 8B). The inheritance pattern of the genes overlapping with inversions for categories 2 and 3 for each dataset is given in Supplementary figure 8C and D. About 40.9% and 50% of the inversions in both categories 2 and 3 regardless of dataset are in AR disease genes (Supplementary figure 8C and D). Autosomal dominant (AD) inheritance is the second most prominent disease gene pattern (16.7% and 33.8%, Supplementary figure 8C and D).

We then performed gene set enrichment analysis with all protein-coding genes in the datasets that intersect with the protein-coding genes following categories 2 and 3. All enriched HPO terms belonging to each category are provided in Supplementary table 7 and 8.

## DISCUSSION

In this study, we reported a case with c.1389+1G>A and NC_000017.11: 75576992_75829587inv in *UNC13D* presenting with an FHL3 phenotype. The inversion in the patient disrupts *UNC13D* following category 2 (Figure 2). The pathogenic inversion is present in heterozygosity in gnomAD (v4.0) in individuals from European Finnish and Admixed American populations (Supplementary Figure 3). To identify more inversions that are likely pathogenic like the one in *UNC13D*, we delved deep into gnomAD inversions. We extensively investigated gnomAD inversions to gain a comprehensive understanding of inversions in an individual genome. 279 rare inversions in gnomAD affect 247 protein-coding genes associated with a phenotype in OMIM based on categories 2 and 3; 254 of them have not been found in the homozygous state and overlap with 106 AR disease genes (Supplementary table 4), similar to the overlap between INV_CHR17_66182818 and *UNC13D*. For instance, 15,736-bp inversion in gnomAD, INV_chr1_04df2580, (https://gnomad.broadinstitute.org/variant/INV_CHR1_04DF2580?dataset=gnomad_sv_r4) disrupts *DPYD* with the breakpoint junctions in intron 12 and intron 8 of *DPYD.* Van Kuilenburg *et al.* has reported a 115,731-bp inversion with breakpoints in intron 8 and intron 12 of *DPYD* in a patient with Dihydropyrimidine dehydrogenase deficiency (OMIM #274270) [37].

Then, we conducted analyses on inversions from diverse datasets. It is important to highlight that these inversions were derived from different sequencing technologies (Table 1). While the inversions in 1KGP and gnomAD were detected using short-read WGS, the inversions reported by Ebert *et al.* and Porubsky *et al.* were identified by long-read WGS and Strand-seq. Strand-seq was shown to be the ideal technology to detect inversions, especially those mediated by large segmental duplications or other genomic repeats which often happen as a result of NAHR; 72% of balanced inversions in Porubsky *et al*. are generated by NAHR [4,22]. In contrast, short-reads are not suitable to identify such inversions, although it can resolve inversions with blunt or microhomology at the breakpoint junctions such as those generated by NHEJ [1]. Therefore, while we expected to detect redundancy among datasets, we also expected to identify unique inversions only identifiable by certain methodologies but invisible to others. While between 11.1% to 49.4% of the inversions in gnomAD overlap with inversions in other datasets, from 21.6% to 76.4% of inversions in Porubsky *et al.* overlap with inversions in other datasets. Strikingly, gnomAD (v4.0) has inversions with a longer length and a higher number of larger inversions (median length of 7.1 kb), which raises the question of whether Mb size inversions, including pericentric ones, are more often generated by NHEJ (Table 1). In fact, we have investigated large inversions detected by karyotyping (8 Mb to 178 Mb) in a diagnostic setting, and found that none of the resolved inversions (13/18 or 72%) are mediated by repeats [1] which has been confirmed by a second more recent study [38]. Besides, it should be taken into account that these inversions were generated by different SV callers, and these tools exhibit different false positive rates [18,39]. These potential false calls may overlap with protein-coding genes in our analysis. Also, redundancies in these datasets will occur due to the same ancestral inversions being reported from distinct individuals while identified by distinct technologies, due to analysis of similar samples or due to the incorporation of entire datasets into larger ones, *e.g*., DGV incorporates 1KGP phase 3 (Supplementary table 2).

Next, we examined whether the inversions in all datasets disrupt human protein-coding genes by classifying inversion-gene intersections into three different categories (Figure 1). The majority of the overlaps in all datasets except 1KGP are from category 1 (Figure 3) which is consistent with the small inversion sizes in 1KGP (Supplementary Figure 5, Table 1) but also indicates that inversions in 1KGP often have both breakpoints within genes which potentially can lead to truncated transcripts subjected to nonsense mediated decay (NMD) or to exon skipping. The results also show that most of the inversions intersecting with protein-coding genes in the other datasets are longer than the gene length. Besides, 97.1% of intersections in gnomAD belong to category 1, consistent with gnomAD presenting longer inversions compared to other datasets (Table 1). Next, we focused on the protein-coding genes that are associated with a phenotype in OMIM disrupted by inversions. Upon examining the genes overlapping with the inversions in categories 2 and 3, we found that most genes intersecting with inversions across all datasets belong to the AR group, while AD disease genes are the second most prominent group. (Figure 4B and Figure 4C). Inversions that disrupt AD disease genes can also be particularly noteworthy, as they might introduce genomic instability in these regions, potentially leading to the formation of other SVs [16].

**Figure 4.**
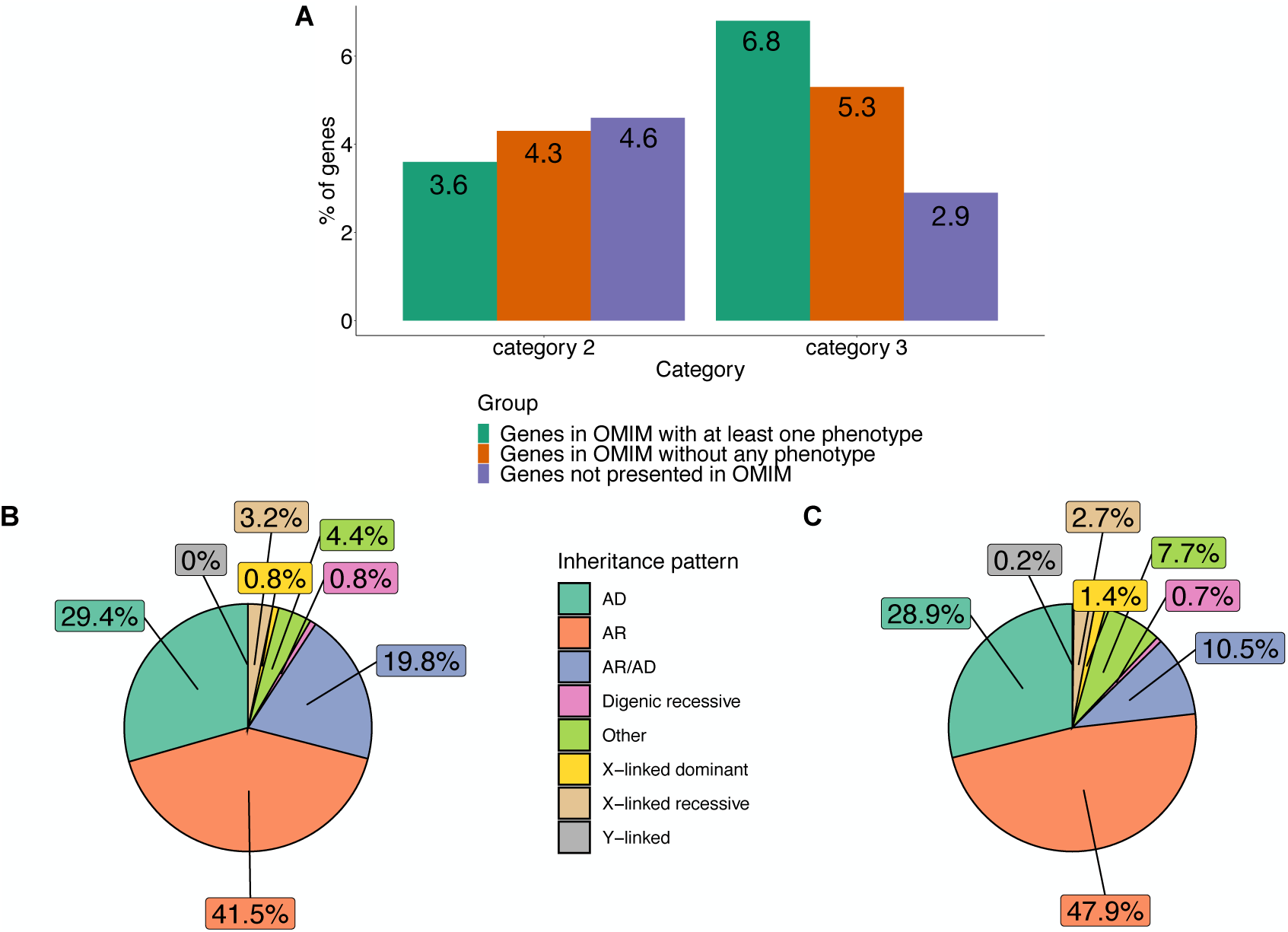
(A) Number of OMIM phenotype-related genes (protein-coding) overlapping with the inversions in all datasets in category 2 and 3. (B) Inheritance pattern of the genes overlapping with all inversions in category 2. (C) Inheritance pattern of the genes overlapping with all inversions in category 3.

The number of inversions involving protein-coding genes associated with one or more phenotypes is markedly distinct in each dataset, with gnomAD and DGV showing a higher overlap rate with OMIM phenotype-related genes than other datasets (Supplementary Figure 8). We observed that the genes disrupted by inversions in categories 2 and 3 are associated with both Mendelian disorders, such as Spinocerebellar ataxia 31 (OMIM #619422), and complex disease traits, such as susceptibility to autism (OMIM #618830).

We further performed gene set enrichment analysis on the genes interrupted by inversions in categories 2 and 3. All enriched HPO terms except Autosomal dominant inheritance (HP:0000006) for category 3 are statistically insignificant (Supplementary table 7 and 8). This result might be expected since we used diverse genes that overlap inversions in the whole genome. Nevertheless, we still report the list of HPO terms enriched by the genes disrupted by inversions to be able to gain an insight into these genes and their related phenotypes.

Finally, sequencing technologies, including short-read WGS, long-read WGS, Strand-seq, and optical mapping, have significantly contributed to the discovery of inversions. Publicly accessible datasets using these technologies are important resources that may facilitate discoveries of pathogenic inversions underlying various disease traits. This study sheds light on the possible impact of the inversions in these datasets on revealing disease phenotypes.

## DATA AVAILABILITY

Gnomad SV data: https://gnomad.broadinstitute.org/downloads

DGV SV data: http://dgv.tcag.ca/dgv/docs/GRCh38_hg38_variants_2020-02-25.txt

1KGP SV data: https://www.internationalgenome.org/data-portal/data-collection/30x-grch38 The updated callset to the original release of the inversions reported by Ebert *et al*. [22]: http://ftp.1000genomes.ebi.ac.uk/vol1/ftp/data_collections/HGSVC2/release/v2.0/integrated_ callset/

GENCODE v46: https://genome.ucsc.edu/cgi-bin/hgTables

OMIM gene list: https://www.omim.org/downloads

Orphanet gene list: https://www.orphadata.com/genes/

## CODE AVAILABILITY

The script for data analysis in this manuscript is available at https://github.com/Carvalho-Lab/Tugce_INV/tree/main.

## ACKNOWLEDGEMENTS

We thank the patient and family for participation in this study.

## AUTHOR CONTRIBUTIONS

Conceptualization: CMBC, ZCA; Data Analysis: TBY and JDB; Funding acquisition: CMBC; Clinical data: IKC; Supervision: CMBC and ZCA; Writing, review, and editing: TBY, MYL, JDB, US, IKC, ZCA, and CMBC. All authors have read and approved the final manuscript.

## ETHICAL APPROVAL

This study is approved by the Baylor College of Medicine (BCM) Institutional Review Board and WIRB for the Pacific Northwest Research Institute (IRB Protocol #H-47127/20202158).

## SUPPLEMENTARY INFORMATION

Supplementary Figure 1. Barplot of the transcript types of the genes in GENCODE v46.

Supplementary Figure 2. IGV visualization of the detected variants in SEA110. (A) Illustration of which mapped reads correspond to each junction, reads mapping to jct2 are the reads highlighted in red with soft clipping extending to green, while reads highlighted in green with soft clipping extending into red map to junction 1. (B) Manual phasing of c.1389+1G>A to the non-inverted haplotype. Black boxes highlight SNPs that are represented unique to the SNV haplotype. The dashed black light indicates a read that extends past jct2 from the inversion and contains c.1389+1G>A.

Supplementary Figure 3. The *UNC13D* inversion in gnomAD.

Supplementary Figure 4. Rare and common inversions in gnomAD. (A) 99% of inversions in gnomAD v4.0 are rare with <0.5% frequency. (B) Number of genes intersecting with common inversions in gnomAD v4.0 based on category 2 and 3. (C) Number of genes intersecting with rare inversions in gnomAD v4.0 based on category 2 and 3.

Supplementary Figure 5. The log2 transformed length plot of the inversion sizes in the datasets.

Supplementary Figure 6. Plot of number of common and dataset-specific inversions in the datasets.

Supplementary Figure 7. Comparison of inversions in each dataset based on overlapping at least 50% of sequence length.

Supplementary Figure 8. The protein-coding genes that are related to a phenotype in OMIM overlapping with inversions. (A) The percentage of the OMIM phenotype-related genes overlapping inversions in category 2. (B) The percentage of the OMIM phenotype-related genes overlapping inversions in category 3. (C) The pie charts of inheritance patterns of genes overlapping with inversions in each dataset based on category 2. (D) The pie charts of inheritance patterns of genes overlapping with inversions in each dataset based on category 3.

Supplementary Table 1. Primer sets used in the study.

Supplementary Table 2. References of DGV inversions.

Supplementary Table 3. Rare gnomAD inversions-genes intersections in category 2 and 3.

Supplementary Table 4. Rare gnomAD inversions with homozygous frequency 0 - OMIM AR genes intersections in category 2 and 3.

Supplementary Table 5. Inversion-gene intersections from all datasets in category 2.

Supplementary Table 6. Inversion-gene intersections from all datasets in category 3.

Supplementary Table 7. Enriched HPO terms for the protein-coding genes intersecting with the inversions in all datasets in category 2.

Supplementary Table 8. Enriched HPO terms for the protein-coding genes intersecting with the inversions in all datasets in category 3.

## REFERENCES

1. Pettersson M, Grochowski CM, Wincent J, Eisfeldt J, Breman AM, Cheung SW, et al. Cytogenetically visible inversions are formed by multiple molecular mechanisms. Human Mutation. 2020;41:1979–98.

2. Flores M, Morales L, Gonzaga-Jauregui C, Domínguez-Vidaña R, Zepeda C, Yañez O, et al. Recurrent DNA inversion rearrangements in the human genome. Proc Natl Acad Sci USA. 2007;104:6099–106.

3. Kidd JM, Cooper GM, Donahue WF, Hayden HS, Sampas N, Graves T, et al. Mapping and sequencing of structural variation from eight human genomes. Nature. 2008;453:56–64.

4. Porubsky D, Höps W, Ashraf H, Hsieh P, Rodriguez-Martin B, Yilmaz F, et al. Recurrent inversion polymorphisms in humans associate with genetic instability and genomic disorders. Cell. 2022;185:1986–2005

5. Carvalho CMB, Lupski JR. Mechanisms underlying structural variant formation in genomic disorders. Nat Rev Genet. 2016;17:224–38.

6. Puig M, Casillas S, Villatoro S, Cáceres M. Human inversions and their functional consequences. Brief Funct Genomics. 2015;14:369–79.

7. Mor-Shaked H, Paz-Ebstein E, Basal A, Ben-Haim S, Grobe H, Heymann S, et al. Levodopa-responsive dystonia caused by biallelic *PRKN* exon inversion invisible to exome sequencing. Brain Communications. 2021;3:fcab197.

8. Jones ML, Murden SL, Brooks C, Maloney V, Manning RA, Gilmour KC, et al. Disruption of AP3B1by a chromosome 5 inversion: a new disease mechanism in Hermansky-Pudlak syndrome type 2. BMC Medical Genetics. 2013;14:42.

9. Meeths M, Chiang SCC, Wood SM, Entesarian M, Schlums H, Bang B, et al. Familial hemophagocytic lymphohistiocytosis type 3 (FHL3) caused by deep intronic mutation and inversion in UNC13D. Blood. 2011;118:5783–93.

10. Qian Y, Johnson JA, Connor JA, Valencia CA, Barasa N, Schubert J, et al. The 253-kb inversion and deep intronic mutations in *UNC13D* are present in North American patients with familial hemophagocytic lymphohistiocytosis 3. Pediatric Blood & Cancer. 2014;61:1034–40.

11. de Jong S, Chepelev I, Janson E, Strengman E, van den Berg LH, Veldink JH, et al. Common inversion polymorphism at 17q21.31 affects expression of multiple genes in tissue-specific manner. BMC Genomics. 2012;13:458.

12. Pilbrow AP, Lewis KA, Perrin MH, Sweet WE, Moravec CS, Tang WHW, et al. Cardiac CRFR1 Expression Is Elevated in Human Heart Failure and Modulated by Genetic Variation and Alternative Splicing. Endocrinology. 2016;157:4865–74.

13. González JR, Ruiz-Arenas C, Cáceres A, Morán I, López-Sánchez M, Alonso L, et al. Polymorphic Inversions Underlie the Shared Genetic Susceptibility of Obesity-Related Diseases. The American Journal of Human Genetics. 2020;106:846–58.

14. Ruiz-Arenas C, Cáceres A, Moreno V, González JR. Common polymorphic inversions at 17q21.31 and 8p23.1 associate with cancer prognosis. Hum Genomics. 2019;13:57.

15. Nomura T, Suzuki S, Miyauchi T, Takeda M, Shinkuma S, Fujita Y, et al. Chromosomal inversions as a hidden disease-modifying factor for somatic recombination phenotypes. JCI Insight. 2018;3:e97595.

16. Osborne LR, Li M, Pober B, Chitayat D, Bodurtha J, Mandel A, et al. A 1.5 million–base pair inversion polymorphism in families with Williams-Beuren syndrome. Nat Genet. 2001;29:321–5.

17. Vicente-Salvador D, Puig M, Gayà-Vidal M, Pacheco S, Giner-Delgado C, Noguera I, et al. Detailed analysis of inversions predicted between two human genomes: errors, real polymorphisms, and their origin and population distribution. Human Molecular Genetics. 2017;26:567–81.

18. Chaisson MJP, Sanders AD, Zhao X, Malhotra A, Porubsky D, Rausch T, et al. Multi-platform discovery of haplotype-resolved structural variation in human genomes. Nat Commun. 2019;10:1784.

19. Cameron DL, Di Stefano L, Papenfuss AT. Comprehensive evaluation and characterisation of short read general-purpose structural variant calling software. Nat Commun. 2019;10:3240.

20. Falconer E, Hills M, Naumann U, Poon SSS, Chavez EA, Sanders AD, et al. DNA template strand sequencing of single-cells maps genomic rearrangements at high resolution. Nat Methods. 2012;9:1107–12.

21. Lam ET, Hastie A, Lin C, Ehrlich D, Das SK, Austin MD, et al. Genome mapping on nanochannel arrays for structural variation analysis and sequence assembly. Nat Biotechnol. 2012;30:771–6.

22. Ebert P, Audano PA, Zhu Q, Rodriguez-Martin B, Porubsky D, Bonder MJ, et al. Haplotype-resolved diverse human genomes and integrated analysis of structural variation. Science. 2021;372:eabf7117.

23. Collins RL, Brand H, Karczewski KJ, Zhao X, Alföldi J, Francioli LC, et al. A structural variation reference for medical and population genetics. Nature. 2020;581:444–51.

24. MacDonald JR, Ziman R, Yuen RKC, Feuk L, Scherer SW. The Database of Genomic Variants: a curated collection of structural variation in the human genome. Nucl Acids Res. 2014;42:D986–92.

25. Byrska-Bishop M, Evani US, Zhao X, Basile AO, Abel HJ, Regier AA, et al. High-coverage whole-genome sequencing of the expanded 1000 Genomes Project cohort including 602 trios. Cell BABABAB. 2022;185:3426–3440.e19.

26. Henter J-I, Horne A, Aricó M, Egeler RM, Filipovich AH, Imashuku S, et al. HLH-2004: Diagnostic and therapeutic guidelines for hemophagocytic lymphohistiocytosis. Pediatr Blood Cancer. 2007;48:124–31.

27. Li H. Minimap2: pairwise alignment for nucleotide sequences. Bioinformatics. 2018;34:3094–100.

28. Zheng Z, Li S, Su J, Leung AW-S, Lam T-W, Luo R. Symphonizing pileup and full-alignment for deep learning-based long-read variant calling. Nat Comput Sci. 2022;2:797–803.

29. Martin M, Patterson M, Garg S, Fischer SO, Pisanti N, Klau GW, et al. WhatsHap: fast and accurate read-based phasing [Internet]. bioRxiv; 2016 [cited 2024 Apr 23]. p. 085050. Available from: https://www.biorxiv.org/content/10.1101/085050v2

30. Haeussler M, Zweig AS, Tyner C, Speir ML, Rosenbloom KR, Raney BJ, et al. The UCSC Genome Browser database: 2019 update. Nucleic Acids Research. 2019;47:D853–8.

31. Amberger JS, Bocchini CA, Scott AF, Hamosh A. OMIM.org: leveraging knowledge across phenotype–gene relationships. Nucleic Acids Research. 2019;47:D1038–43.

32. Quinlan AR, Hall IM. BEDTools: a flexible suite of utilities for comparing genomic features. Bioinformatics. 2010;26:841–2.

33. Chen EY, Tan CM, Kou Y, Duan Q, Wang Z, Meirelles GV, et al. Enrichr: interactive and collaborative HTML5 gene list enrichment analysis tool. BMC Bioinformatics. 2013;14:128.

34. R Core Team. R: A Language and Environment for Statistical Computing [Internet]. Vienna, Austria: R Foundation for Statistical Computing; 2023. Available from: https://www.R-project.org/

35. Hadley Wickham. ggplot2: Elegant Graphics for Data Analysis [Internet]. Springer-Verlag New York; 2016. Available from: https://ggplot2.tidyverse.org

36. Conway JR, Lex A, Gehlenborg N. UpSetR: an R package for the visualization of intersecting sets and their properties. Bioinformatics. 2017;33:2938–40.

37. Van Kuilenburg ABP, Tarailo-Graovac M, Meijer J, Drogemoller B, Vockley J, Maurer D, et al. Genome sequencing reveals a novel genetic mechanism underlying dihydropyrimidine dehydrogenase deficiency: A novel missense variant c.1700G>A and a large intragenic inversion in *DPYD* spanning intron 8 to intron 12. Human Mutation. 2018;39:947–53.

38. Bilgrav Saether K, Eisfeldt J, Bengtsson J, Lun MY, Grochowski CM, Mahmoud M, et al. Mind the gap: the relevance of the genome reference to resolve rare and pathogenic inversions. medRxiv. 2024;2024.04.22.24305780.

39. Kosugi S, Momozawa Y, Liu X, Terao C, Kubo M, Kamatani Y. Comprehensive evaluation of structural variation detection algorithms for whole genome sequencing. Genome Biol. 2019;20:117.

