## Supplementary figures for "Investigation of a Pathogenic Inversion in *UNC13D* and Comprehensive Analysis of Chromosomal Inversions Across Diverse Datasets"

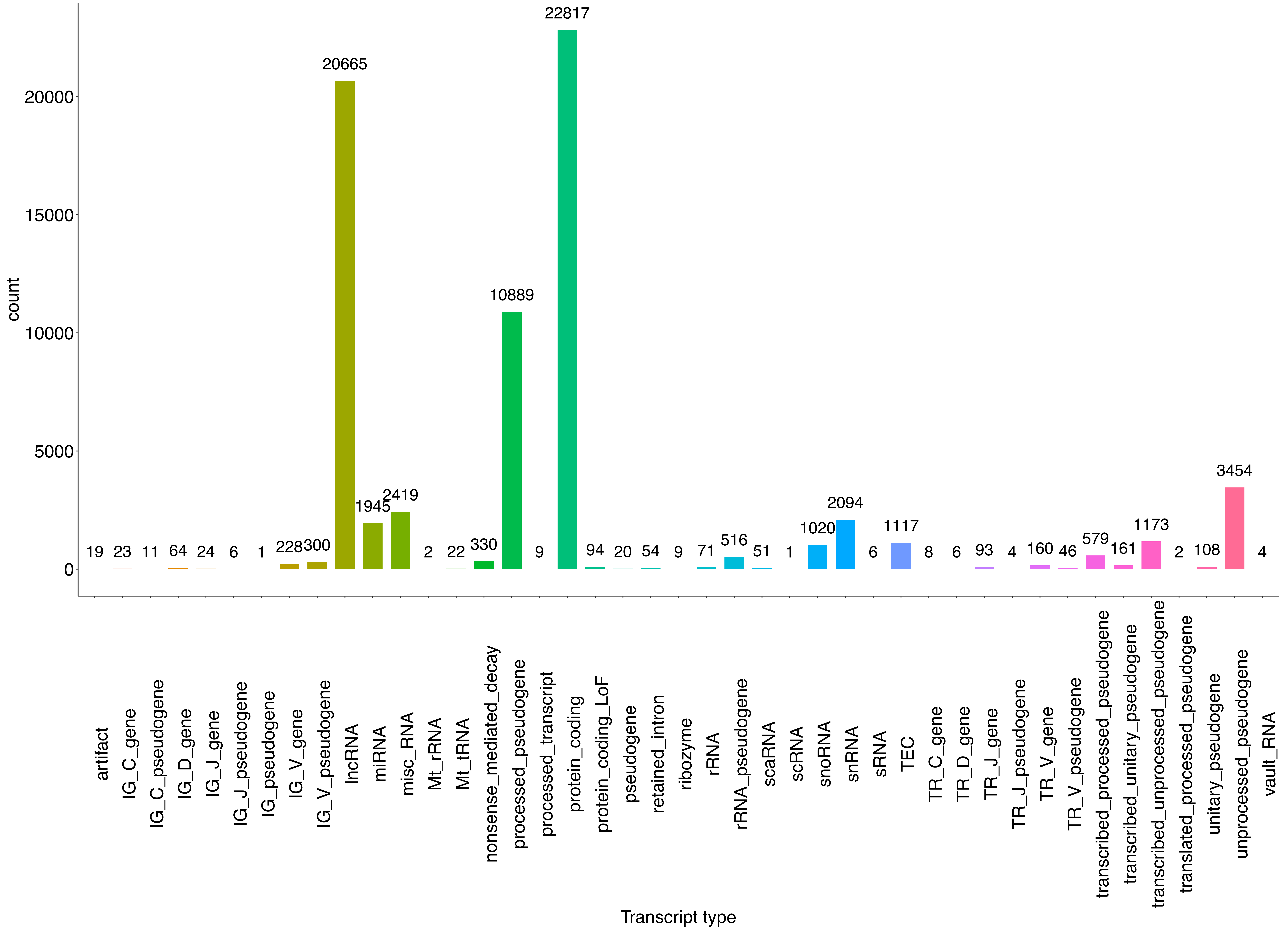


**Supplementary figure 1**

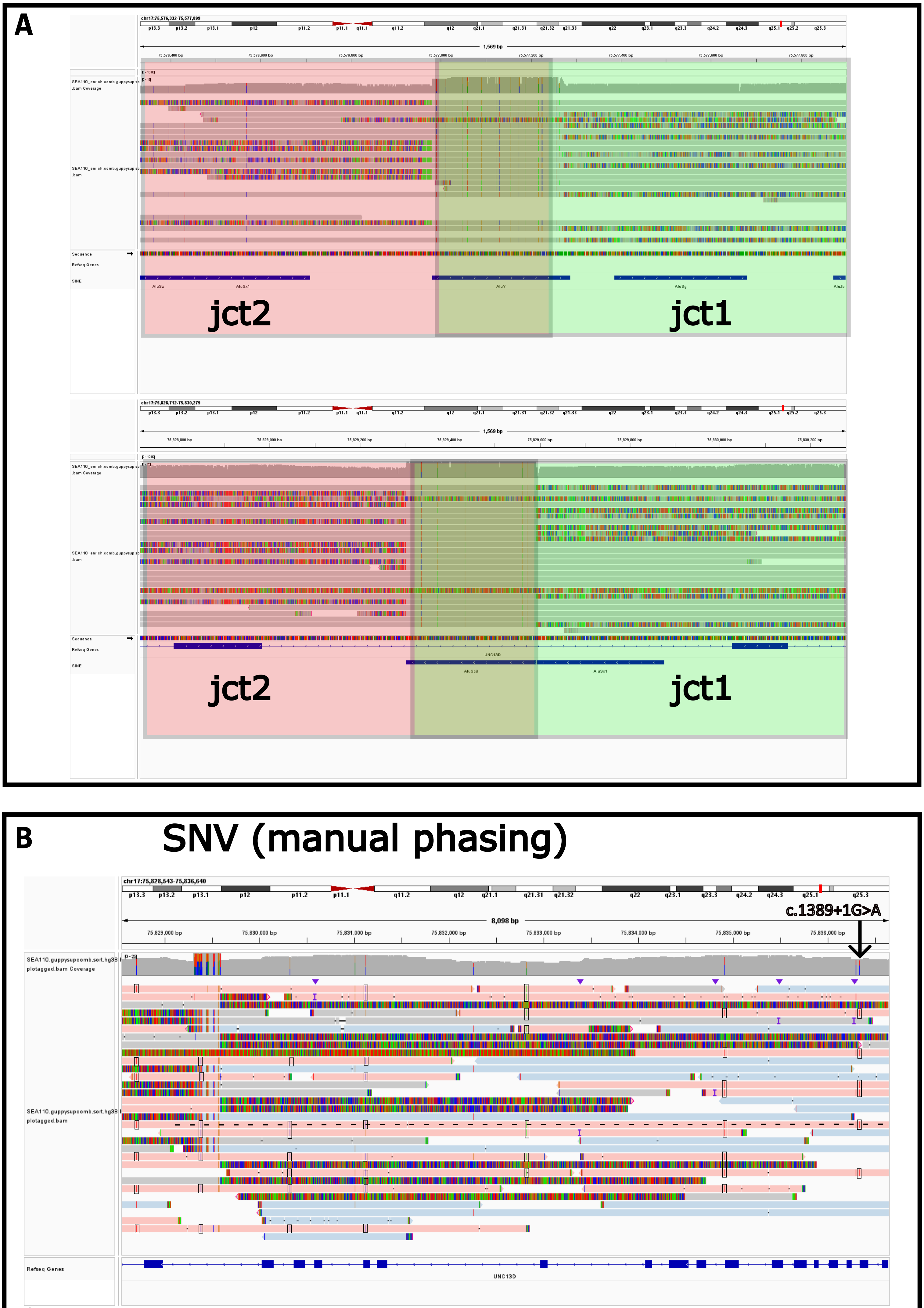


Supplementary figure 2

**Filter** Pass
**Allele Count** 8
**Allele Number** 126092
**Allele Frequency** 0.00006345
**Quality score** 1
**Position** 17:75576924-75829482
**Size** 252,558 bp
**Class** inversion
**Evidence** Anomalous paired-end reads
**Algorithms** TEXT NEEDED FOR ALGORITHM LABEL "rescan", Manta

### External Resources

- UCSC

### Feedback

Report an issue with this variant

### Genetic Ancestry Group Frequencies

| Genetic Ancestry Group | Allele Count | Allele Number | Number of Homozygotes | Allele Frequency |
|---|---|---|---|---|
| Admixed American | 2 | 12594 | 0 | 0.0001588 |
| European (Finnish) | 1 | 6476 | 0 | 0.0001544 |
| European (non-Finnish) | 5 | 59088 | 0 | 0.00008462 |
| African/African American | 0 | 33816 | 0 | 0.000 |
| Amish | 0 | 842 | 0 | 0.000 |
| Ashkenazi Jewish | 0 | 3180 | 0 | 0.000 |
| East Asian | 0 | 4054 | 0 | 0.000 |
| Middle Eastern | 0 | 64 | 0 | 0.000 |
| Remaining | 0 | 1572 | 0 | 0.000 |
| South Asian | 0 | 4406 | 0 | 0.000 |
| XX | 3 | 65800 | 0 | 0.00004559 |
| XY | 5 | 60292 | 0 | 0.00008293 |
| **Total** | **8** | **126092** | **0** | **0.00006345** |

**Supplementary figure 3**

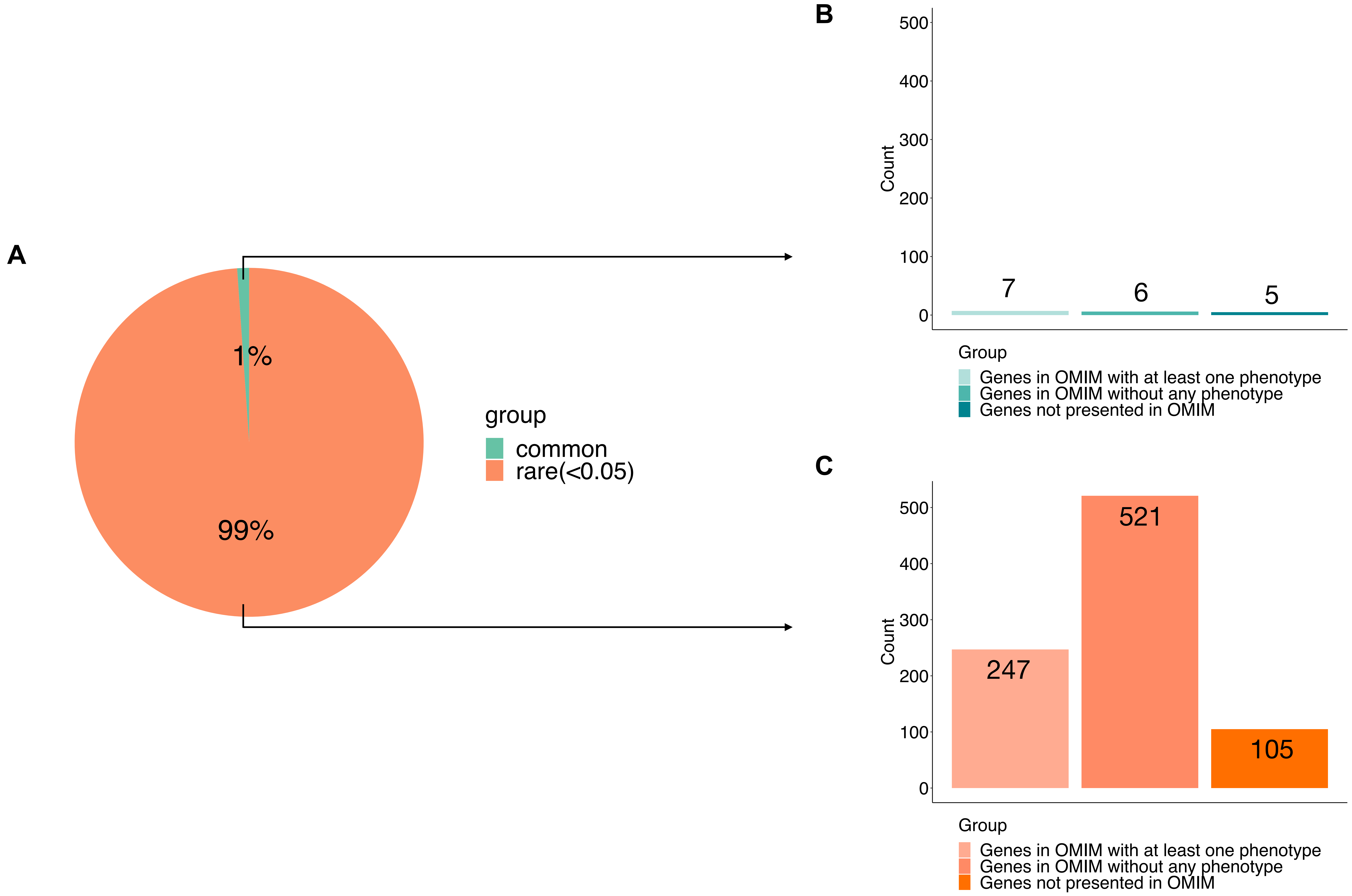


**Supplementary figure 4**

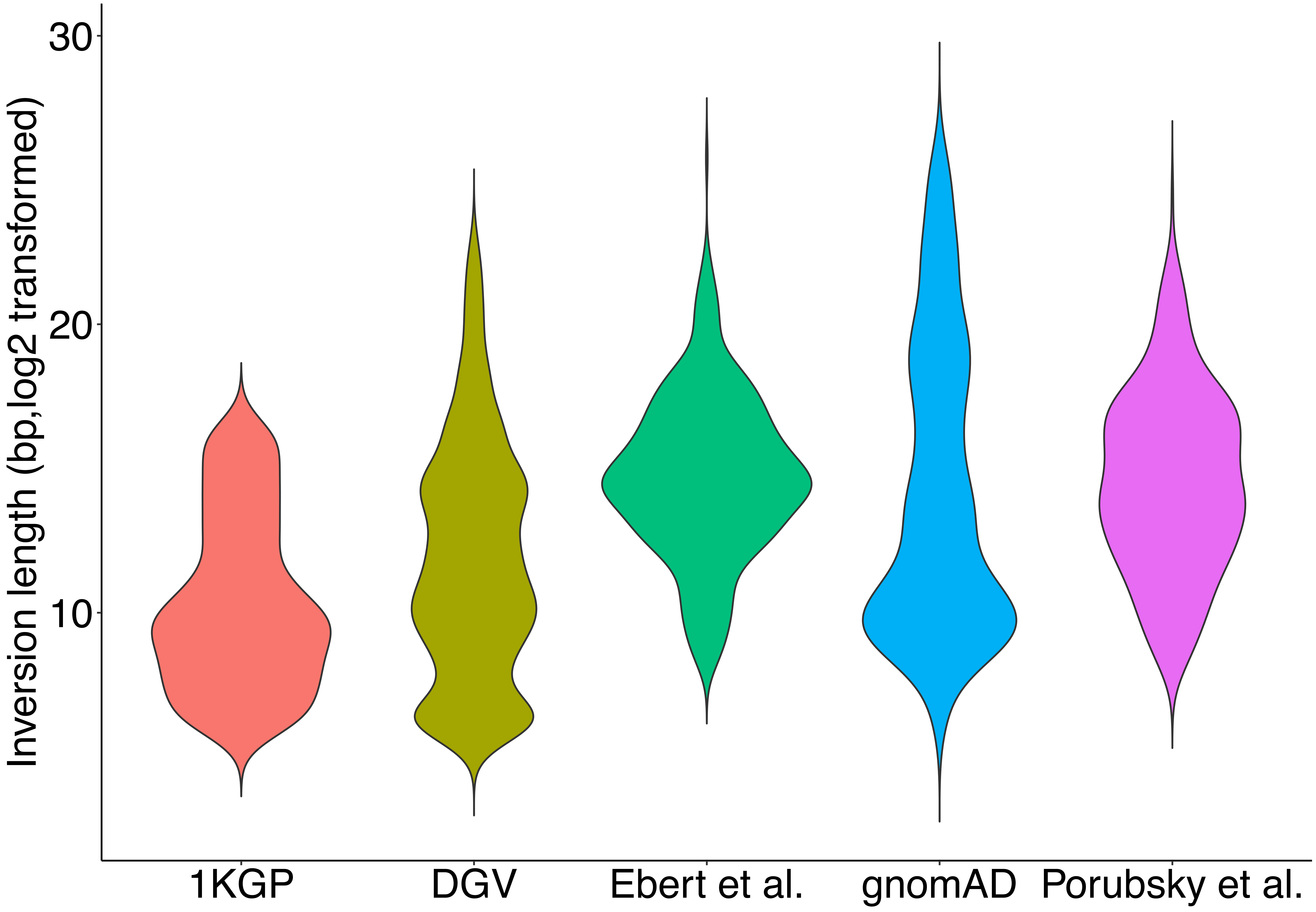


**Supplementary figure 5**

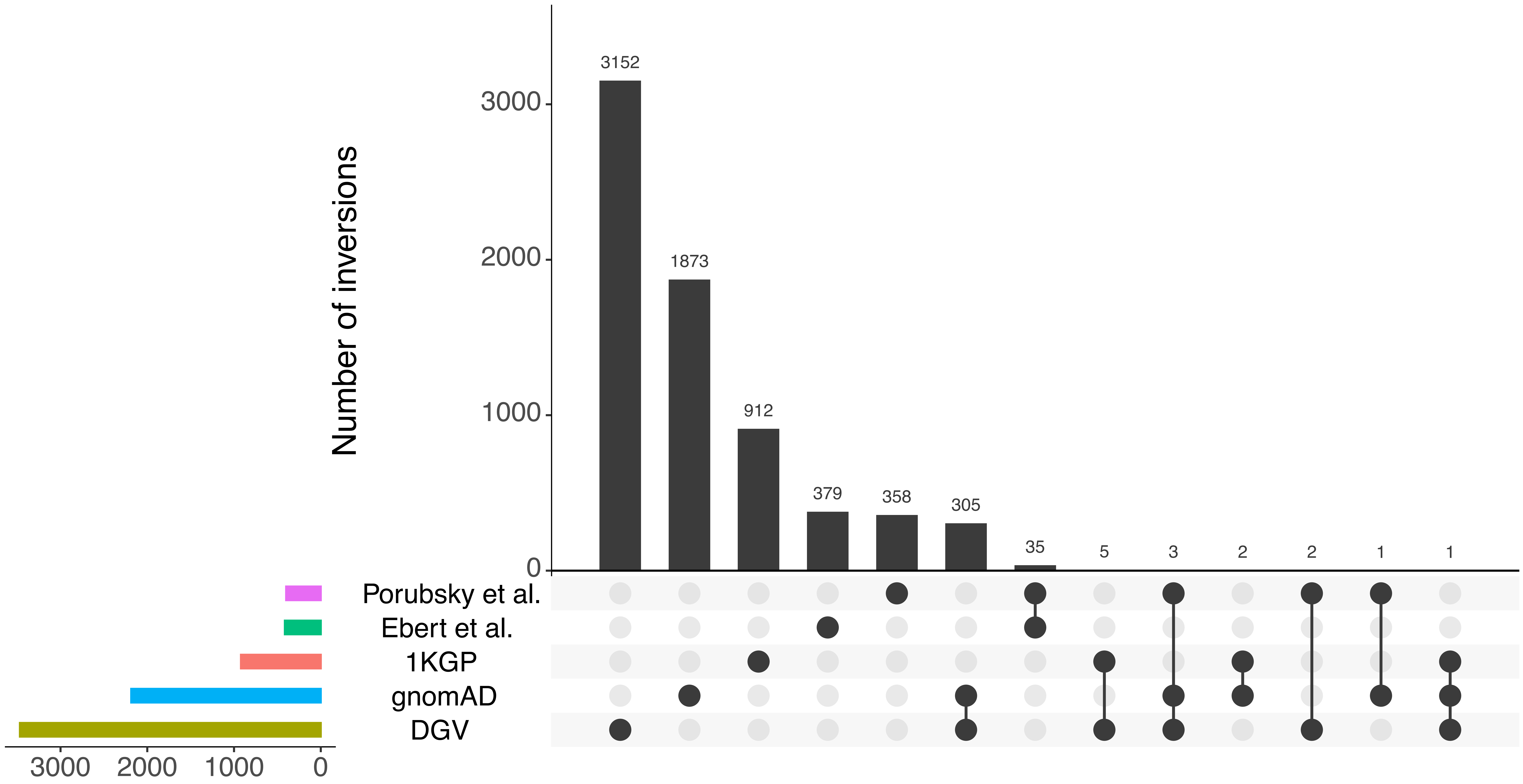


**Supplementary figure 6**

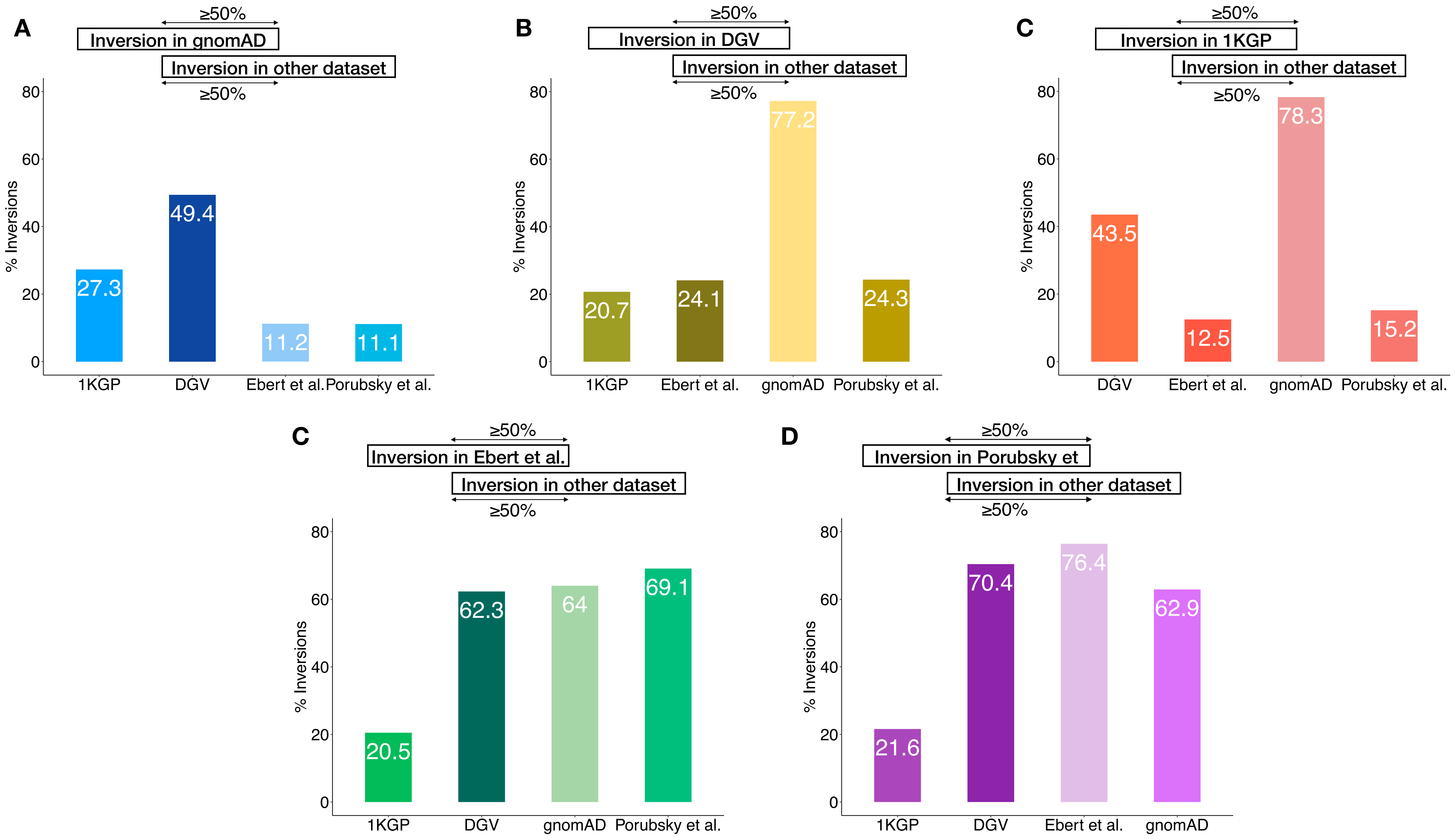


**Supplementary figure 7**

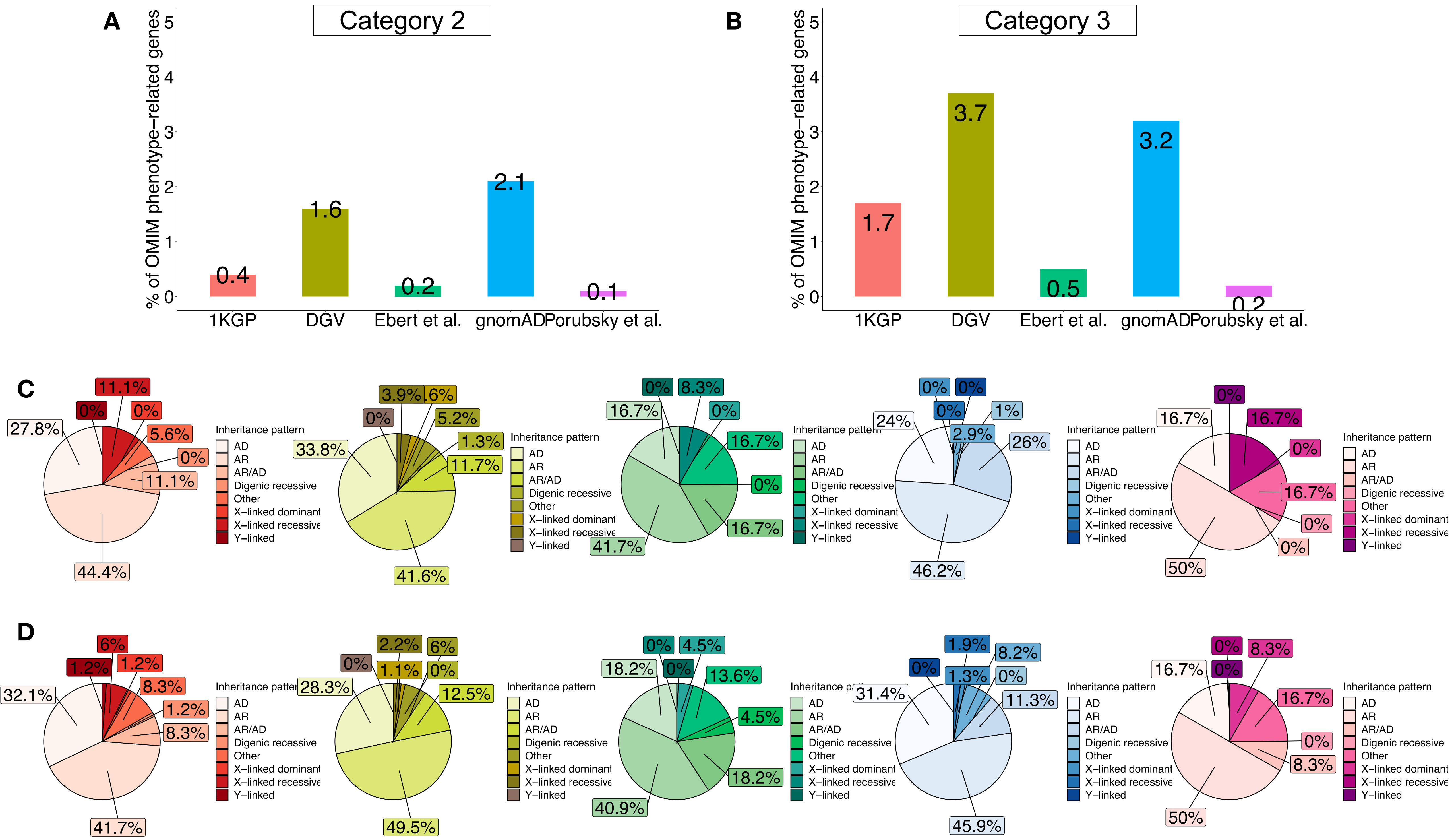


**Supplementary figure 8**
